# Vaccine effectiveness of the maternal RSVpre-F vaccine against severe disease in infants in Scotland, UK: national population-based case-control and cohort analyses

**DOI:** 10.1101/2025.08.01.25332515

**Authors:** Isobel McLachlan, Chris Robertson, Kirsty E Morrison, Ross McQueenie, Safraj Shahul Hameed, Cheryl Gibbons, Rachael Wood, Rachel Merrick, Louisa Pollock, Antonia Ho, Ting Shi, Thomas C Williams, Aziz Sheikh, Jim McMenamin, Sam Ghebrehewet, Kimberly Marsh

## Abstract

**Background:** Respiratory syncytial virus (RSV) is a leading cause of infant hospitalisation, particularly in those under six months. In response, Scotland introduced a maternal RSVpreF vaccination programme in 2024, offering the vaccine from 28 weeks gestation. While clinical trials demonstrate high efficacy, real-world evidence is needed to assess vaccine effectiveness (VE) to inform policy and programme delivery.

**Methods:** We conducted a retrospective, nested case–control study within the full population of infants aged ≤90 days in Scotland (n=27,552) during the 2024/25 peak RSV season, using routinely collected clinical data to estimate the VE of maternal RSV vaccination against RSV-related lower respiratory tract infection (LRTI) hospitalisations. Cases were infants hospitalised with a LRTI and a positive RSV polymerase chain reaction (PCR) test within 14 days before or two days after admission, between August 12, 2024 and March 31, 2025. Each case was matched to 10 controls by birth week and gestational age. VE was estimated using adjusted conditional logistic regression, calculated as 100 × (1−odds ratio).

**Findings:** During the study period, 13,878 (50.4%) pregnant women received the vaccine. 350 infants aged ≤90 days were hospitalised with RSV, with 3,471 matched controls included in the analysis. Among cases, 60 (17.1%) were born to vaccinated mothers, compared to 1,704 (49.1%) among controls. The median week of gestation at vaccination was 30 weeks (interquartile range 28-33 weeks). VE against RSV-associated LRTI hospitalisation was 82.9% (95%CI: 75.9-87.8). VE remained high among preterm infants (<37 weeks: 89.2%, 95%CI: 52.2-97.6). Sub-optimal immunisation (<14 days between vaccination and birth) did not confer protection (29.6%, 95%CI: −19.6–58.6). Vaccination averted 228 (95%CI: 197-252) RSV-related LRTI hospitalisations in infants aged ≤90 days.

**Interpretation:** In this first national, population-based study, we provide robust evidence that maternal RSV vaccination provides substantive protection against RSV-related hospitalisation in infants ≤90 days, including those born pre-term. Maternal RSV vaccination programmes need to be scaled up globally at pace.

*Evidence before this study:* Respiratory syncytial virus (RSV) is a leading cause of lower respiratory tract infections (LRTI) in infants and young children globally, contributing significantly to hospitalisations and healthcare burden each year. While most infections are mild, RSV can lead to severe disease in infants, particularly those under six months of age, in preterm infants and in children with underlying medical conditions. Recently, the UK approved use of the Abrysvo® Pre-F RSV (henceforth RSVpre-F) vaccine among pregnant women to protect infants against severe outcomes of RSV, representing a major shift in the public health management of this pathogen. Given that the RSVpre-F vaccine has only been delivered in a small number of countries to date, there are limited studies showing real-world vaccine effectiveness (VE) of RSV vaccines given to pregnant women to protect infants. We searched PubMed originally on July 1, 2025, then performed a follow-up search on July 29, 2025, using the search terms ((((RSV) OR (respiratory syncytial)) AND (vaccine effectiveness)) AND (maternal)) AND (pregnancy). The literature search was restricted to literature produced between June 21, 2023 (the date that the RSV vaccine was recommended in the USA) and July 29, 2025. There were no language restrictions in our searches. Our searches returned 34 papers. Of these, two were qualitative studies, five were editorials, two were modelling studies, four examined cost-effectiveness of the vaccine, nine were literature reviews, and nine investigated vaccine efficacy rather than effectiveness. Under randomised clinical trial conditions, the reported vaccine efficacy against hospitalisation ranged from 65.5% to 100%. The remaining three papers from our literature review described RSV maternal VE in Argentina and the UK, using case-control studies similar to our analyses but conducted in selected medical centres rather than across the full population. The first paper by Marc et al. reported VE against RSV-related hospitalisation of 78.6% (95% confidence interval (CI): 62.1-78.9) in those aged 0-3 months and 71.3% (95% CI: 53.5-82.3) in those aged 0-6 months. The second by Gentile et al. reported VE of 78.7% (95% CI: 51.4-90.7) in those aged 0–3 months and 68.2% (95% CI: 33.1-84.9) in those aged 0-6 months. In both studies, RSV vaccine was administered to pregnant women between 32 and 36 weeks of gestation and used multi-centre test negative case-control designs. The final paper by Williams et al., a multi-centre study carried out over Scotland and England, reported a VE of 72% for infants whose mothers were vaccinated more than 14 days before delivery.

*Added value of this study:* We use a nested case-control methodology to estimate VE against RSV-related hospitalisation among all Scottish infants aged ≤90 days, born between August 12, 2024 and March 31, 2025. Vaccines were given to pregnant women from 28 weeks’ gestation-with a median week of gestion of 30 weeks. This is earlier than the Argentinian studies, where the vaccine was administered between 32-36 weeks, and vaccine efficacy trials, where the median gestational week of vaccination was 31 weeks. Rather than a multi-centre approach, we used population-wide routinely collected clinical data to examine all hospitalisations of infants in Scotland with an RSV positive PCR test and coded with LRTI. Individual-level mother-baby data linked through national clinical, vaccination, and laboratory data systems were used to estimate VE. Our results showed high population-level VEs, with an 82.9% (95% CI: 75.9–87.8) reduction in RSV-related LRTI hospitalisations among infants ≤90 days whose mothers were vaccinated compared to unvaccinated. From this, we estimated that 228 (95% CI: 197-252) RSV-related LRTI hospitalisations were averted in babies in Scotland across the study period. To test the robustness of findings, we undertook a secondary retrospective matched cohort design, showing a similar estimate of VE against RSV-related LRTI hospitalisations at 81.0% (95% CI: 68.6-88.5). Further, we were able to show that the vaccine was effective against RSV-related LRTI hospitalisations in preterm infants (<37 weeks; 89.2%, 95%CI: 52.2-97.6), and that sub-optimal immunisation (<14 days between vaccination and birth) did not confer protection against hospitalisation (29.6%, 95%CI: −19.6–58.6). This study represents the first real-world evidence of maternal RSV VE in a national, population-based cohort of pregnant women vaccinated from 28 weeks’ gestation. Further, this study is the first to report the effects of sub-optimal immunisation in infants from mothers who were vaccinated less than two weeks before giving birth, the first to evidence protection for preterm babies, who are typically at greater risk of RSV-related hospitalisations than full term babies, and the first to provide an estimate of RSV-related hospitalisations averted. Our study also uses a separate validation approach using a retrospective matched cohort design to overcome potential bias from unmeasured confounding associated with infant factors.

*Implications of all the available evidence:* RSV vaccination was effective against LRTI hospitalisations, including in babies born prematurely. In summary, we provide robust evidence of the substantial protection afforded by maternal RSV vaccination offered from 28 weeks’ gestation against RSV-related hospitalisation in infants.

## Introduction

Respiratory syncytial virus (RSV) is a leading cause of acute respiratory illness globally, with the majority of children infected by age two.^1,2^ While typically mild, infants under six months, especially those born prematurely and those with chronic conditions, are at increased risk of severe complications, including bronchiolitis and pneumonia.^3,4^ Severe early-life RSV infection is associated with long-term respiratory morbidity, including recurrent wheezing, asthma, impaired lung function, and chronic respiratory disease into adulthood.^5^

Globally, in 2019, RSV was estimated to cause approximately 33.0 million episodes of acute lower respiratory tract infection (LRTI) in children under five, leading to 3.6 million hospital admissions and up to 101,400 deaths, nearly half of which occurred in infants under six months.^1^ Between 2000 to 2011, 2.1% of children in Scotland were hospitalised with RSV by age two, most within their first year.^6^ RSV accounted for 6.2% of all hospital bed days and 6.9% of intensive care unit bed days in infants, highlighting a substantial burden in young children and on healthcare resources, especially during the winter.^6^ Like much of the Northern Hemisphere, Scotland experiences a distinct RSV season, with most cases occurring between October and February.^7^

To reduce the burden of RSV infection in young infants in the UK, the Joint Committee on Vaccination and Immunisation (JCVI) recommended the implementation of an RSV immunisation programme.^8^ In response to the JCVI’s recommendation, the UK and Scottish governments implemented a maternal RSV immunisation programme, selecting the maternal vaccine as the preferred strategy to confer passive immunity to infants through transplacental antibody transfer.^9^

Scotland launched its maternal RSV vaccination programme on August 12, 2024, offering a single dose of RSVpreF (Abrysvo; Pfizer, New York, NY, USA) to pregnant women from 28 weeks’ gestation.^10^ A catch-up campaign was also implemented for those who were between 29 weeks and full term when the vaccine was introduced. The vaccine is provided by National Health Service (NHS) Scotland, year-round and free of charge as part of routine antenatal care. This routine maternal immunisation programme is distinct from, and in addition to, a longstanding selective programme recommending RSV monoclonal antibody immunisation for infants born very or extremely prematurely (less than 32 weeks) and infants and young children who are particularly susceptible to severe RSV infection.^11^

Clinical trials have demonstrated high efficacy of RSVpreF in preventing RSV-associated LRTI in infants, with limited early real-world studies supporting effectiveness.^12–14^ High quality real-world VE studies remain critical for generating robust, context-specific evidence across varied populations and implementation strategies. This evidence not only strengthens public confidence in immunisation programmes, but also informs policy decisions, particularly in countries evaluating infant RSV prevention strategies amid the availability of both maternal vaccines and monoclonal antibody products. We present the first population-based real-world estimates of VE against RSV-related hospitalisations – and hospitalisations averted – from all hospitals in Scotland in infants ≤90 days, following the rollout of Scotland’s national maternal RSV vaccination programme from 28 weeks’ gestation.

## Methods

### Study design and population

A pre-specified statistical analysis plan is provided in Supplementary Material 1. We conducted national population-based case-control and cohort analyses to estimate VE of the maternal RSV vaccine against RSV-related LRTI hospitalisations in infants aged ≤90 days. The study population comprised all singleton live births born in Scotland between August 12, 2024, and March 31, 2025. Pregnancies with multiple fetuses were excluded to avoid bias from shared genetics and environment, which could reduce exposure differences between cases and controls. Individuals entered the study population at birth and were censored at RSV positive test not related to hospitalisation, transfer out of GP records (driven predominantly by individuals leaving the country), age >90 days, death or the end of the study period, whichever occurred first.

### Data sources

The datasets and data items used in these analyses are shown in Supplementary Table 1. Datasets were accessed through the Public Health Scotland’s (PHS) Scottish Infectious Respiratory Surveillance Platform (SIRSP), which is a deduplicated, clinical register of all patients in NHS Scotland. Patients within the Scottish NHS system had a unique patient Community Health Index (CHI) number. The SIRSP was pseudonymised with individuals assigned a respiratory identifier to enable linkage to datasets within SIRSP.

Singleton live births were identified using Scottish Linked Pregnancy and Baby Dataset (SLiPBD), a population-based cohort of all recognised pregnancies and births in Scotland from 2000 to present.^15^ Records of live births in SLiPBD contain unique patient identifiers for both the mother and the baby, allowing linkage or records relating to mother-baby pairs. RSV vaccination data were linked to SLiPBD using the mother’s respiratory identifier and all other datasets were linked using the infant’s respiratory identifier.

Records of RSV vaccination received during pregnancy, including those administered in Scotland and to Scottish residents vaccinated elsewhere, were obtained from the National Clinical Data Store (NCDS).^16^ RSV positive tests, obtained from Electronic Communication of Surveillance Scotland (ECOSS),^17^ and hospital admissions, obtained from Scottish Morbidity Records (SMR01),^18^ were used to determine RSV-related hospital admissions among babies in the study population. ECOSS is a PHS surveillance system that receives laboratory test results from all Scottish NHS laboratories. SMR01 contains episode-level data on inpatient and day-case discharges from acute specialties (excluding neonatology) in Scottish hospitals. General Practitioner (GP) records were used to identify infants born in Scotland that were transferred out and therefore lost to follow up. Infant deaths were identified from SLiPBD and from the National Records of Scotland (NRS) deaths dataset, which contained all registrations of deaths to NRS.^19^

### Case definition: RSV-related respiratory hospitalisation

Cases were infants at aged ≤90 days that had an RSV-related LRTI respiratory hospitalisations. An RSV-related LRTI hospitalisation was defined as an emergency admission recorded with a LRTI diagnosis (Supplementary Table 2) listed in any diagnostic position as the reason for hospital admission, and an RSV positive −PCR test with a specimen date occurring within 14 days before or two days after hospital admission. Sensitivity analysis was conducted using a broader case definition of an RSV-related respiratory hospitalisation at aged ≤90 days. An RSV-related respiratory hospitalisation was defined as an emergency admission recorded with a respiratory diagnosis (Supplementary Table 2) listed in any diagnostic position as the reason for hospital admission, and an RSV positive PCR test with a specimen date occurring within 14 days before or two days after hospital admission.

### Controls and matching

Cases were matched to controls at the time of RSV-related LRTI (or respiratory, sensitivity analysis) hospitalisation using a 1:10 case: control ratio. Eligible controls were selected from the study population and had no previous RSV positive PCR test or RSV-related hospitalisation at the time of matching. Controls were matched on International Organization for Standardization (ISO) week of birth and gestational age (i.e. number of weeks’ gestation delivered at) at birth to account for RSV seasonality and age-related disease susceptibility. Weekly number of births were low for pre-term babies (born at <37 weeks gestation). Gestational age was therefore categorised as <32 weeks gestation, 32-34 weeks, 35-36 weeks and in individual weeks from 37 weeks onward to increase the number of complete case-control sets. A case was only a case once and could not later become a control, however controls could later become cases. The ccwc() function in Epi (version 2.59), R (version 4.4.2) was used for matching cases and controls.

### Exposure: maternal RSV vaccination

Maternal RSV vaccine status was determined by linking RSV vaccination records of the mothers to infants in study population. Infants were classified as vaccinated if the maternal RSV vaccine was given during pregnancy and more than 14 days before delivery. Birth more than 14 days after vaccination of a pregnant woman has been shown to be sufficient time for transplacental transfer of antibodies to newborns with some antibody transfer when birth occurred within two weeks of vaccination.^20^ Where a vaccinated mother gave birth within 0-14 days of receiving the RSV vaccine (either due to pre-term birth, delayed uptake of the vaccine offer, or due to being part of the catch up campaign), infants were classified as having sub-optimal immunisation. Infants were classified as unvaccinated if their mothers did not receive the RSV vaccine during pregnancy.

### Statistical analysis

Odds ratios (ORs) and 95% confidence intervals (CIs) comparing individuals whose mothers had been vaccinated with those whose mothers had not been vaccinated among cases and controls were calculated using unadjusted and adjusted conditional logistic regression models. The adjusted model accounted for maternal age at end of pregnancy, birthweight, maternal ethnicity, parity, baby’s sex, maternal Scottish Index of Multiple Deprivation (SIMD) at end of pregnancy, and maternal smoking at antenatal booking. The adjusted and unadjusted conditional logistic regression equations used to calculate OR estimates are shown in Supplementary Equation 1. VE for infants ≤90 days and by gestational age was calculated using the following equation:

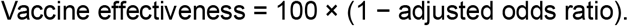

Sub-analyses were carried out calculating adjusted odds ratios (aORs) of vaccinated term (≥37 weeks gestation) and vaccinated pre-term (<37 weeks gestation) births (by including an interaction between vaccination status and gestation at birth (term or pre-term) in the model) and aORs comparing vaccinated infants and infants with sub-optimal immunisation.

To assess the robustness of the RSV maternal VE results derived from the nested case-control design, we conducted a sensitivity analysis using a retrospective matched cohort design. Using this methodology, mothers were matched before birth, where factors associated with the baby are unknown and therefore cannot influence the matching. In the matched cohort design, pregnant women vaccinated between August 12, 2024 to March 31, 2025 were matched 1:1 to unvaccinated pregnant women at the date of vaccination by age and week of gestation. A 1:1 matching ratio was chosen as it was statically efficient and avoided the additional complexity and diminishing returns of higher ratios. Follow up began on the infant’s date of birth or September 30, 2024, whichever occurred later, until date of first RSV-related hospitalisation, end of study or infant death. Analysis was conducted using a Cox proportional hazards regression, based upon calendar time, adjusted for SIMD, parity, maternal ethnicity, maternal smoking status, birthweight and baby sex, and stratified by matched pair. Matched pairs were removed from analysis if the unvaccinated match was vaccinated before birth (N=2,550), or either of the matched pair had a multiple birth (N=184).

Hospitalisations averted following vaccination was calculated using Supplementary Equation 2 and 3 (adapted from Savinkina et al.).^21^

Analyses were conducted using R/RStudio, version 4.4.2. Packages: Epi (version 2.59).

### Ethical considerations and permissions

This study was conducted using routinely collected, pseudo-anonymised health data held by Public Health Scotland. Access to general practice data was via Public Health Data (Infectious Respiratory Diseases) (Scotland) Directions 2024. All analyses were conducted within a secure data environment in compliance with PHS data governance and information security protocols. No patient-level identifiers were accessed for these analyses.

### Reporting

A STROBE/RECORD checklist was completed for this study and is shown in Supplementary Table 7. A pre-specified analysis plan is available within the supplementary data.

### Role of the funding source

This study received no specific funding.

## Results

### Characteristics of the study population

During the study period, 13,878 (50.4%) pregnant women received the vaccine. Median gestational age at maternal vaccination was 29 weeks (IQR: 26-32 weeks) (Table 1). A total of 1,094 (7.0%) infants were born before 37 weeks and therefore considered preterm. Among infants born at term, 47.5% (n=12,165) were to mothers who had received the RSV vaccine >14 days before delivery compared to 31.9% (n=619) in pre-term infants (<37 weeks gestation). Notably, most infants with sub-optimal immunisation were born at term (970 of 1,094, 88.7%).

**Table 1:** Characteristics of pregnant women and infants with and without RSV-related LRTI hospital admissions aged ≤90 days, in Scotland, August 12, 2024 to March 31, 2025.

| Characteristic | RSV-related admission at ≤90 days (Cases) | No RSV-related admission | No RSV-related admission (Matched Controls)* | Case/Control p-value <sup>a</sup> |
| --- | --- | --- | --- | --- |
| Number | 350 | 27,202 | 3,471 |  |
| **Infant characteristics** |  |  |  |  |
| **Age at RSV-related LRTI admission (days)** | 52 (34–68) | - | - | - |
| **Birthweight (g), median (IQR)** | 3,422.5 (2,990-3,745) | 3,420 (3,080-3,745) | 3,350 (2,991-3,690) | 0.16<sup>b</sup> |
| **Gestational age at vaccination (weeks)** | 29 (26-32) | 31 (29-34) | 30 (28-33) | <0.05<sup>b</sup> |
| **Gestational age at birth** |  |  |  |  |
| Gestational age at birth (weeks), median (IQR) | 39 (38-40) | 39 (38-40) | 39 (38-40) | 0.82<sup>b</sup> |
| Gestational age at birth: <37 weeks | 41 (11.7%) | 1,899 (7.0%) | 381 (11.0%) | <0.05 |
| Gestational age at birth: ≥37 weeks | 309 (88.3%) | 25,303 (93.0%) | 3,090 (89.0%) |  |
| **Sex** |  |  |  |  |
| Female | 151 (43.1%) | 13,262 (48.8%) | 1,658 (47.8%) | 0.10 |
| Male | 199 (56.9%) | 13,939 (51.2%) | 1,813 (52.2%) |  |
| Unknown | 0 (0%) | 1 (<0.1%) | 0 (0%) |  |
| **Maternal characteristics** |  |  |  |  |
| **Vaccination Status** |  |  |  |  |
| Time between vaccination and delivery among those who received a RSV vaccine during pregnancy (days), median (IQR) | 30 (10-52) | 52 (33-70) | 41.5 (24-59) | <0.05<sup>b</sup> |
| Unvaccinated | 290 (82.9%) | 13,384 (49.2%) | 1,767 (50.9%) | <0.05 |
| Sub-optimal immunisation | 18 (5.1%) | 1,076 (4.0%) | 201 (5.8%) |  |
| Vaccinated | 42 (12.0%) | 12,742 (46.8%) | 1,503 (43.3%) |  |
| **Smoking status** |  |  |  |  |
| Ex-smoker | 40 (11.4%) | 3,196 (11.7%) | 437 (12.6%) | <0.05 |
| Non-smoker | 247 (70.6%) | 21,567 (79.3%) | 2,733 (78.7%) |  |
| Smoker | 62 (17.7%) | 2,321 (8.5%) | 285 (8.2%) |  |
| Unknown | 1 (0.3%) | 118 (0.4%) | 16 (0.5%) |  |
| **Parity** |  |  |  |  |
| 0 | 65 (18.6%) | 12,098 (44.5%) | 1,458 (42.0%) | <0.05 |
| 1 | 152 (43.4%) | 8,592 (31.6%) | 1,136 (32.7%) |  |
| 2+ | 117 (33.4%) | 4,687 (17.2%) | 679 (19.6%) |  |
| Unknown | 16 (4.6%) | 1,825 (6.7%) | 198 (5.7%) |  |
| **Maternal age** |  |  |  |  |
| (years), median (IQR) | 31 (27-34) | 31 (27-35) | 31 (27-35) | <0.05<sup>b</sup> |
| <25 years | 58 (16.6%) | 3,707 (13.6%) | 449 (12.9%) | <0.05 |
| 25-29 years | 101 (28.9%) | 6,860 (25.2%) | 862 (24.8%) |  |
| 30-34 years | 116 (33.1%) | 9,620 (35.4%) | 1,227 (35.4%) |  |
| 35+ years | 75 (21.4%) | 7,014 (25.8%) | 933 (26.9%) |  |
| Unknown | 0 (0%) | 1 (<0.1%) | 0 (0%) |  |
| **SIMD (at end of pregnancy)** |  |  |  |  |
| 1 (most deprived) | 122 (34.9%) | 6,571 (24.2%) | 842 (24.3%) | <0.05 |
| 2 | 71 (20.3%) | 5,666 (20.8%) | 711 (20.5%) |  |
| 3 | 58 (16.6%) | 5,052 (18.6%) | 673 (19.4%) |  |
| 4 | 57 (16.3%) | 5,512 (20.3%) | 667 (19.2%) |  |
| 5 (least deprived) | 42 (12.0%) | 4,365 (16.0%) | 573 (16.5%) |  |
| Unknown | 0 (0%) | 36 (0.1%) | 5 (0.1%) |  |
| **Ethnicity** |  |  |  |  |
| Black/ Caribbean/ African | 8 (2.3%) | 1,162 (4.3%) | 150 (4.3%) | 0.10 |
| South Asian | 14 (4.0%) | 1,639 (6.0%) | 217 (6.3%) |  |
| White | 306 (87.2%) | 22,448 (82.5%) | 2,863 (82.5%) |  |
| Other or mixed ethnicity | 19 (5.4%) | 1,535 (5.6%) | 189 (5.4%) |  |
| Unknown/ missing | 3 (0.9%) | 418 (1.5%) | 52 (1.5%) |  |

\* 32 controls later became cases and therefore some individuals may be counted in the table more than once. <sup>a</sup> Pearson's Chi-squared test <sup>b</sup> Mann-Whitney test

We identified 350 RSV-related LRTI hospital admissions among infants aged ≤90 days during the study period (August 12, 2024 to March 31, 2025). Admission trends across the study period are shown in Figure 1. High numbers of early vaccinations were driven by a (front-loaded) catch-up programme, where all pregnant women over 28 weeks of gestation on August 12, 2024 were offered vaccination at the start of the campaign. RSV-related hospital admissions peaked in late November 2024, consistent with reported RSV incidence reaching very high levels (defined as incidence greater than 327.7 per 100,000) for those aged under one year in weeks 47 and 49.^7^

**Figure 1:**
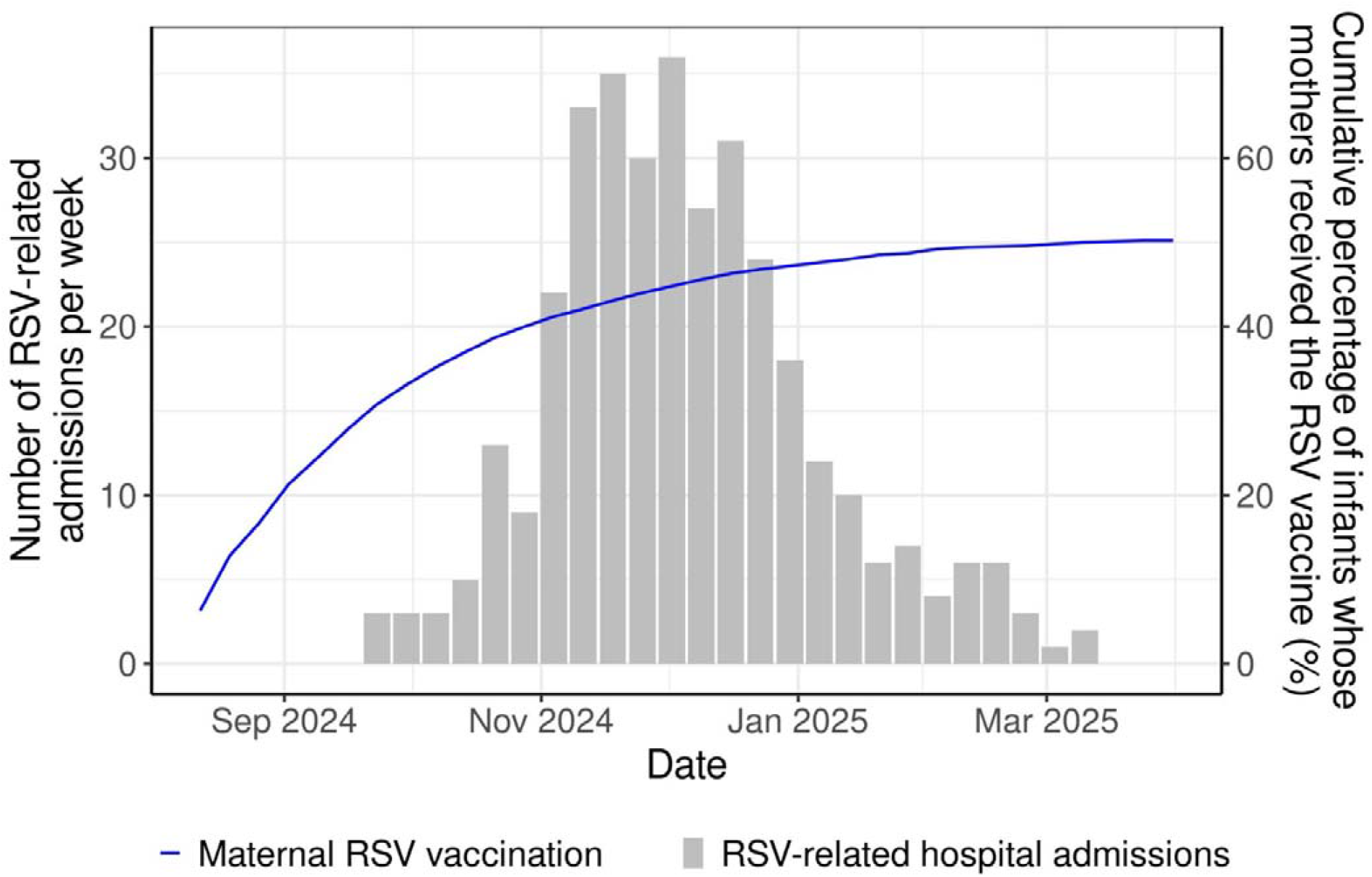
Maternal RSV vaccination and RSV-related LRTI hospital admissions among infants aged ≤90 days in Scotland, August 12, 2024 to March 31, 2025.

### Characteristics of cases and controls

350 RSV-related LRTI hospital admissions (cases) were identified and matched to 3,471 controls. Five of the 350 cases could not be matched to 10 controls due to insufficient availability of eligible controls meeting the matching criteria. Among the controls, 32 later became cases. The median age of cases at RSV-related LRTI admission was 52 days (IQR 34-68 days) (Table 1). The median gestational age at birth was 39 weeks (IQR 38-40 weeks) (Table 2) for both cases and control. Compared to controls, cases were significantly more likely to be born to younger mothers, pregnant women who smoked, had previous births, or lived in areas of higher deprivation (p < 0.05). Additionally, a significantly higher proportion of cases were unvaccinated against RSV (82.9%, 290/350) compared to controls (50.9%, 1,767/3,471; p < 0.05; Table 2). Median gestational age at vaccination was 31 weeks among cases (IQR: 29-34 weeks) and 30 weeks IQR: 28-33 weeks) among controls (Table 1).

### Vaccine effectiveness and impact

Among infants aged ≤90 days, maternal RSV vaccination (>14 days before delivery) was associated with a VE against RSV-related LRTI hospital admissions of 82.9% (95% CI: 75.9-87.8) (Figure 2, Supplementary Table 3). This translated to 228 (95% CI: 197-252) RSV-related LRTI hospitalisations averted in babies aged ≤90 days over the study period.

**Figure 2:**
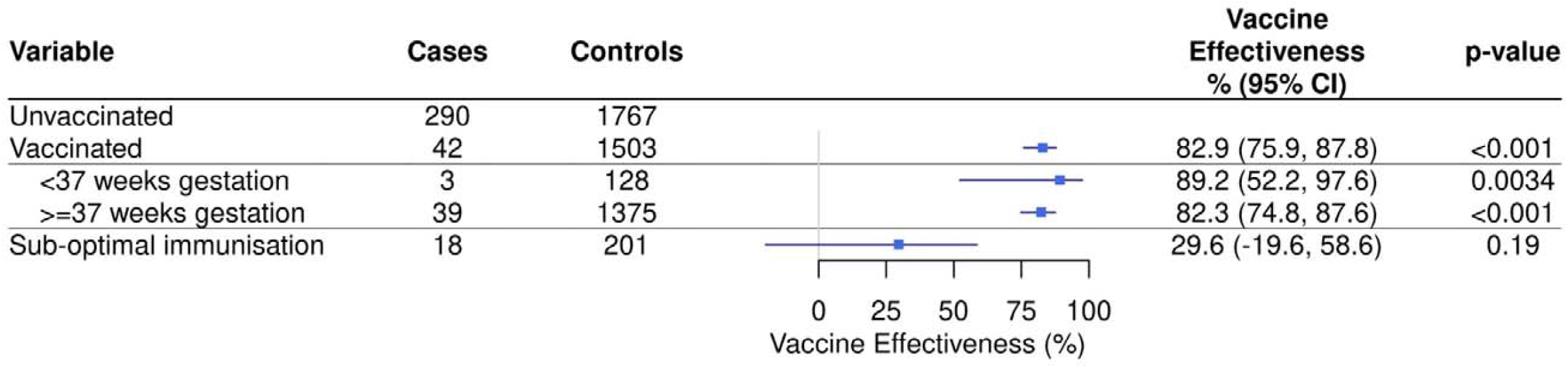
Forest plot of maternal RSV VE (%) against RSV-related LRTI hospitalisation in infants aged ≤90 days.

### Subgroup analysis

Our results showed significant protection for pre-term infants, with a VE of 89.2% (95% CI: 52.2-97.6) against RSV-related LRTI admissions. VE among infants with sub-optimal immunisation was 29.6% (95% CI: −19.6-58.6) against RSV-related LRTI admissions, signalling no clear protective effect within this group.

### Retrospective matched cohort design

To test the robustness of results from the nested case/control design, these analyses were repeated using a retrospective matched cohort design. There were 231 RSV-related LRTI admissions in infants aged ≤90 days identified in the retrospective matched cohort design. Compared to the nested case-control design this analysis was based on fewer RSV-related LRTI admissions, with most of the missing admission in unvaccinated babies whose mothers were not selected in the matching process. RSV-related LRTI admissions for these matched pairs are shown in Supplementary Table 5. Using a Cox’s proportional hazards model (Supplementary Table 6), we found that the adjusted VE for vaccinated pregnant women was 81.0% (95% CI: 68.6-88.5). No significant VE was observed in pregnant women with sub-optimally immunisation (−28.6%, 95% CI: −224.5-49.0).

### RSV-related admissions with any respiratory diagnosis

Results were consistent using a broader definition of RSV-related admission with a respiratory (as opposed to an LRTI only) diagnosis (Supplementary Table 4; vaccinated VE 80.4% (95% CI: 73.1-85.7), vaccinated term VE 79.4% (95% CI: 71.4-85.1) pre-term VE 89.3% (95% CI: 62.8-96.9) infants and sub-optimal immunisation VE 34.8% (95% CI: −8.2-60.7)).

## Discussion

This national, population-based study provides the first real-world evidence of maternal RSV VE against RSV related hospitalisation in infants at a population level. The RSVpre-F vaccine showed significant protection against RSV LRTI related admissions in infants aged ≤90 days, reducing the risk of hospitalisation in the vaccinated group by 82.9% (95% CI: 75.9-87.8) compared to the unvaccinated group. From this, we estimated that 228 (95% CI: 197-252) RSV-related LRTI hospitalisations were averted over the study period. Further, we have shown for the first time that the RSV vaccine provided significant protection against RSV related hospitalisation in preterm babies < 37 weeks gestation (VE 89.2%, 95%CI: 52.2-97.6), and that there was no clear protective VE against RSV related hospitalisations in infants whose mothers were vaccinated fewer than 14 days of giving birth (VE 29.6%, 95% CI: −19.6-58.6).

A key strength of this study was the availability of high-quality, linked electronic health records on vaccination, pregnancy outcomes, hospitalisation, and pathogen testing covering the maternal and infant population. Scotland’s robust clinical data systems comprehensively capture all recognised pregnancies, births, hospitalisations, and vaccination records, enabling accurate identification of maternal vaccination status and eligible mother-infant pairs. These high-quality data minimise bias and support reliable, population-level insights. Engagement with antenatal care services in the UK is also consistently high; however, disparities persist in the timing of initial appointments, with women from more deprived areas less likely to attend within the recommended gestational window.^22^

Limitations of this study include its restriction to a single RSV season, which precludes analysis of the duration of protection and potential variation in VE across different seasons. Although we examined outcomes in preterm infants, numbers were insufficient to differentiate between very and moderately preterm births. Future studies are planned to investigate both of these effects by carrying out cumulative analyses over multiple seasons, and by combining data with other UK nations to increase sample sizes. A further limitation is that individual-level prescribing data for Palivizumab were not available; however, we were able to establish that the number of doses provided to the hospitals were relatively small. Hospitals were issued with 1,354 doses to be administered over the course of 5 months, meaning the maximum number of infants treated with Palivizumab in Scotland – assuming full doses are administered - was 270. It should be further noted that this is restricted specifically to preterm babies or those with at-risk conditions,^11^ meaning it is unlikely to have significantly contributed to averted hospitalisations in our population, and doses may be administered outside of the study period. Finally, estimates of hospitalisations averted did not account for wider population-level transmission dynamics following the introduction of the vaccination programme. Although more sophisticated modelling would be needed to refine these estimates, the impact is likely to be small given the targeted nature of the programme and current uptake levels.

Our findings are consistent with existing published studies, including a meta-analysis of three maternal RSV vaccine trials reporting pooled efficacy of 81.9% (95% CI: 56.8–92.4) against RSV-related severe LRTI in infants under three months of age.^23^ Findings from two multicentre case–control studies conducted in Argentina during the 2024 RSV season reported similar results. One prospective study reported a VE of 78.7% (95% CI: 51.4–90.7) against RSV-related hospitalisations in infants under three months.^12^ The BERNI study found a VE of 78.6% (95% CI: 62.1–87.9) against LRTI leading to hospitalisation in the same age group.^13^ A UK multi-centre test-negative case-control study reported a VE of 72% (95% CI: 48–85) against RSV-associated ALRI hospitalisation in infants whose mothers were vaccinated more than 14 days before delivery, during the 2024/25 RSV season.^14^ The consistency of these findings across diverse geographical settings and study designs reinforces the robustness of maternal RSV vaccination as an effective strategy to prevent severe RSV disease in early infancy.

Our findings also support the immunogenicity of maternal RSV vaccination from 28 weeks’ gestation – earlier than the 32–36 week window recommended in countries such as Argentina and the USA that presumably aims to minimise any theoretical risk of preterm birth.^12,13.^ This earlier window enables sufficient time for antibody development and transplacental transfer to occur, maximising neonatal protection, particularly for infants born prematurely. Ongoing safety monitoring, however, is also required to investigate as yet inconclusive signals of increased preterm birth. ^24,25^ These investigations are beyond the scope of this study, but future analyses are planned using Scottish data.

Our results also demonstrate protection in preterm infants, a group at higher risk of severe RSV disease.^4^ While maternal vaccination appears effective in this population, further research is needed to assess durability of protection for the infant. This is particularly important given that Scotland has recently approved the use of nirsevimab, a long-acting monoclonal antibody for passive immunisation that offers additional protection as a one-off immunisation against RSV in very premature infants born before 32 weeks as a complement to the maternal vaccination strategies.^26^

We have also shown here that sub-optimal immunisation is not associated with significant protection, reinforcing the importance of timely administration to establish full protection. Immunological evidence supports vaccination at least five weeks before delivery to optimise antibody transfer.^27^ These findings validate Scotland’s policy of offering RSVpreF from 28 weeks’ gestation and highlight the need for clear guidance to ensure timely uptake.

Despite the demonstrated effectiveness, vaccine uptake during the study period was modest at 50.4%, mirroring rates in England, but higher than those in Wales.^28,29^ To optimise the public health impact of maternal RSV vaccination, efforts should focus on increasing uptake well before the early winter peak in RSV rates, since offer of vaccination at 28 weeks can introduce up to a 12-week gap between uptake and birth, when the benefits from passive protection are most needed.

Our findings demonstrate that maternal RSV vaccination provides substantial protection against RSV-associated hospitalisation in infants during the first 90 days of life, a period of heightened vulnerability. The vaccination programme has the potential to avert hundreds of further RSV-associated hospitalisations in infants annually in Scotland if coverage can be improved. Continued efforts to raise awareness, address barriers to access, and support maternal RSV vaccination in routine antenatal care are vital to maximising the full public health impact of this immunisation programme.

## Contributors

CR, KEM, RM, and KM conceived the study. IM, CR, KEM, RM, SSH, CG, RW, RM, LP, AH, TS, TW, AS, JM, SG and KM made substantial contributions to the design of the work. IM and CR performed the analysis of the data, and IM, CR, KEM, SSH, RW, RM, RM, TM, AS and KM contributed towards interpretation of the data. IM, JM, RM and KM produced a first draft of the manuscript. All authors revised the manuscript and approved the final version of the manuscript in advance of submission. All authors confirm the accuracy and completeness of data used here.

## Data Availability

The R code used for statistical analyses in this study is available upon request. Due to the terms of data access, individual-level data used in this analysis cannot be shared publicly.

## Acknowledgements

The authors would like to thank Mike Birnie, Dan McPhail and Nicholas Young for managing data access; at Public Health Scotland, and UK colleagues in the other three nations, including Conall Watson, Jamie Lopez Bernal, Simon Cottrell, Declan Bradley, Nick Andrews, Heather Whitaker, Freja Kirsebom, Mai Berry, Anna Mensah, and Magda Bucholc for helpful discussions on RSV maternal vaccination. Sincere appreciation is also offered for the substantial work undertaken by NHS Health Board staff, midwives and other clinical staff to support programme delivery and data collection.

## Potential conflict of interests

LP and AH have received research grant funding from Pfizer Inc for work separate from this study. All other authors declare no conflict of interests.

## Notes

### Competing Interest Statement

Louisa Pollock and Antonia Ho have received research grant funding from Pfizer Inc for work separate from this study. All other authors declare no conflict of interests.

### Author Declarations

No specific ethnical approval or waivers for approval are required for this work. This study was conducted using routinely collected, pseudo-anonymised health data held by Public Health Scotland. Access to general practice data was via Public Health Data (Infectious Respiratory Diseases) (Scotland) Directions 2024. All analyses were conducted within a secure data environment in compliance with PHS data governance and information security protocols. No patient-level identifiers were accessed for these analyses.

