## Supplementary Material for "Vaccine effectiveness of the maternal RSVpre-F vaccine against severe disease in infants in Scotland, UK: national population-based case-control and cohort analyses"

**Supplementary Equation 1: Unadjusted and adjusted conditional logistic regression equations**

Unadjusted: $clogit(outcome\sim protection+strata\left( Set \right))$

Adjusted: $clogit(outcome\sim protection+age+birthweight+ethnicity+parity+sex+SIMD+smoking+strata\left( Set \right))$

Unadjusted: $clogit(outcome\sim protection*term+strata\left( Set \right))$

Adjusted: $clogit(outcome\sim protection*term+age+birthweight+ethnicity+parity+sex+SIMD+smoking+strata\left( Set \right))$

Where**:**

- Protection: vaccination status (unvaccinated, sub-optimal immunity (birth within 2 weeks of vaccination), vaccinated)
- Age: maternal age at end of pregnancy (<25, 25-29, 30-34, 35+ years)
- Birthweight: birthweight (very low (<1500g), low (1500-2499g) and normal (≥2500g))
- Ethnicity: maternal ethnicity (Black/Caribbean/African, South Asian, White, Other or mixed ethnicity)
- Parity: number of previous deliveries (0, 1, 2+)
- Set: case-control group
- Sex: baby’s sex (female, male)
- SIMD: maternal Scottish Index of Multiple Deprivation (SIMD) at the end of pregnancy (1 (most deprived), 2, 3, 4, 5 (least deprived))
- Smoking: maternal smoking at antenatal booking (ex-smoker, non-smoker, smoker)
- Term: gestational age at birth (pre-term - <37 weeks gestation and term - ≥37 weeks gestation)

Supplementary Equation 2: Calculation for hospitalisations averted following vaccination

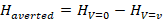

Supplementary Equation 3: Calculation for expected hospitalisations if there had been no vaccination

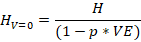

Where:

- *H:* number of hospitalisations,
-
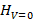
: number of hospitalisations if there had been no vaccination
-
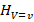
: number of hospitalisations among vaccinated individuals
- *p:* proportion of the population vaccinated,
- VE: vaccine effectiveness

**Supplementary Table 1: List of variables and data sources used in analysis**

|  | **Data Category** | **Data Item** | **Data Source** |
| --- | --- | --- | --- |
| **Exposure** | **Vaccination** | Vaccine | Vaccinations dataset/NCDS |
|  |  | Vaccine date | Vaccinations dataset/NCDS |
| **Outcome** | **Laboratory testing** | RSV PCR test result | ECOSS |
|  |  | Specimen date | ECOSS |
|  | **Mortality** | NRS death | NRS |
|  |  | Infant death flag | SLiPBD |
|  | **Other** | Transfer out | GP |
|  | **Secondary care** | Admission date | SMR01 |
|  |  | Emergency admission flag | SMR01 |
|  |  | Diagnosis | SMR01 |
| **Confounders/**  **covariates** | **Birth** | Birthweight | SLiPBD |
|  |  | End of pregnancy (date) | SLiPBD |
|  |  | Gestational age (at birth) | SLiPBD |
|  |  | Outcome (live births) | SLiPBD |
|  |  | Number births this pregnancy | SLiPBD |
|  |  | Number previous deliveries | SLiPBD |
|  | **Socio-demographics** | Age (mother, at end of pregnancy) | SLiPBD |
|  |  | Ethnicity (mother) | SLiPBD |
|  |  | NHS health board of residence (at booking) | SLiPBD |
|  |  | Sex (baby) | SLiPBD |
|  |  | SIMD (mother, at end of pregnancy) | SLiPBD |
|  | **Other** | Smoking (mother, at antenatal booking) | SLiPBD |

Electronic Communication of Surveillance Scotland (ECOSS), GP records, Scottish Morbidity Records (SMR01), Scottish Linked Pregnancy and Baby Dataset (SLiPBD), National Records of Scotland (NRS), Vaccinations dataset: Data for vaccinations occurring in Scotland, and for Scottish residents vaccinated elsewhere are held in the National Clinical Data Store (NCDS)

**Supplementary Table 2: ICD-10 codes used in RSV-related hospital admission case definition**

| **Definition** | **ICD-10 codes** |
| --- | --- |
| Lower Respiratory Tract Infection (LRTI) diagnosis | J10.0  J11.0  J12  J13  J14  J15  J18  J20  J21  J22  J44.0  J90 |
| Any respiratory diagnosis | J00-J99  U07.1  U07.2 |

**Supplementary Table 3: Maternal RSV vaccine effectiveness (%) against RSV-related LRTI hospitalisation in infants aged ≤90 days.**

| Case definition | Vaccination status | Gestational age at birth | RSV-related admissions (cases) | No RSV-related admission matched controls* | Unadjusted vaccine effectiveness | Adjusted  vaccine effectiveness ^a^ |
| --- | --- | --- | --- | --- | --- | --- |
| **Lower respiratory tract infection (LRTI) ICD-10 code** | **Vaccinated** ^b^ |  | 42 | 1,503 | 84.6% (78.7-88.8%) * | 82.9% (75.9-87.8%) * |
|  |  | **>=37 weeks** | 39 | 1,545 | 84.2% (78.0-88.7%) * | 82.3% (74.8-87.6%) * |
|  |  | **<37 weeks** | 3 | 128 | 87.9% (59.8-96.4%) * | 89.2% (52.2-97.6%) * |
|  | **Sub-optimally immunised** |  | 18 | 201 | 40.7% (3.9-63.4%) p-value = 0.034 | 29.6% (-19.6-58.6%) p-value = 0.19 |

^a^ model adjusts for maternal age, birthweight, maternal ethnicity, parity, baby sex, maternal SIMD and maternal smoking

^b^ Vaccinated includes individuals whose mothers received the RSV vaccine >14 days before delivery

* p-value < 0.01

**Supplementary Table 4: Maternal RSV vaccine effectiveness (%) against any respiratory hospitalisation in infants aged ≤90 days.**

| Case definition | Vaccination status | Gestational age at birth | RSV-related admissions (N=386 cases) | No RSV-related admission matched controls* (N=3,831) | Unadjusted vaccine effectiveness | Adjusted vaccine effectiveness ^a^ |
| --- | --- | --- | --- | --- | --- | --- |
| **Any respiratory ICD-10 code** | **Vaccinated** ^b^ |  | 50 | 1,602 | 82.8% (76.9-87.2%) * | 80.4% (73.1-85.7%) * |
|  |  | **>=37 weeks** | 47 | 1,465 | 82.4% (76.1-87.0%) * | 79.4% (71.4-85.1%) * |
|  |  | **<37 weeks** | 3 | 137 | 87.3% (62.7-95.7%) * | 89.3% (62.8-96.9%) * |
|  | **Sub-optimally immunised** |  | 18 | 221 | 49.0% (17.6-68.5%) p-value = 0.006 | 34.8% ([-8.2]-60.7%) p-value = 0.10 |

^a^ model adjusts for maternal age, birthweight, ethnicity, parity, baby sex, maternal SIMD and maternal smoking

^b^ Vaccinated individuals whose mothers received the RSV vaccine >14 days before delivery

* p-value < 0.01

**Supplementary Table 5: RSV-related LRTI admissions for vaccinated and unvaccinated pregnant women in retrospective matched cohort study**

| **Status** | **Person Years** | **RSV admissions** | **Rate/100** |
| --- | --- | --- | --- |
| Unvaccinated | 1680.2 | 187 | 11.1 |
| All Vaccinated | 1704.5 | 44 | 2.6 |
| Sub-optimal immunity | 184.7 | 17 | 9.2 |
| Vaccinated | 1519.8 | 27 | 1.8 |

**Supplementary Table 6: Maternal RSV vaccine effectiveness (%) against RSV related LRTI hospitalisation in infants aged ≤90 days using a retrospective matched cohort design**

| Analysis type | Vaccination status | Unadjusted | Adjusted ^a^ |
| --- | --- | --- | --- |
| Matched | Sub-optimal immunity | 40.7% (-10.0-68.1%) p-value=0.09 | 44.0% (-14.8-72.7%) p-value=0.11 |
|  | Vaccinated^a^ | 82.9% (73.8-88.8%) * | 81.0% (68.6-88.5%) * |
| Unmatched | Sub-optimal immunity | 26.8% (-21.5-55.9%) p-value=0.22 | 21.7% (-31.0-53.2%) p-value=0.35 |
|  | Vaccinated^a^ | 84.0% (76.0-89.3%) * | 79.5% (68.9-86.6%) * |

* p-value < 0.01

^a^Vaccinated individuals whose mothers received the RSV vaccine >14 days before delivery

**Supplementary Table 7: STROBE/RECORD checklist**

|  | **Item No.** | **STROBE items** | **Location in manuscript where items are reported** | **RECORD items** | **Location in manuscript where items are reported** |
| --- | --- | --- | --- | --- | --- |
| **Title and abstract** | | | | | |
|  | 1 | (a) Indicate the study’s design with a commonly used term in the title or the abstract (b) Provide in the abstract an informative and balanced summary of what was done and what was found | Page 1 | RECORD 1.1: The type of data used should be specified in the title or abstract. When possible, the name of the databases used should be included.  RECORD 1.2: If applicable, the geographic region and timeframe within which the study took place should be reported in the title or abstract.  RECORD 1.3: If linkage between databases was conducted for the study, this should be clearly stated in the title or abstract. | Pages 1-2  Pages 1-2  Page 6 |
| **Introduction** | | | | | |
| Background rationale | 2 | Explain the scientific background and rationale for the  investigation being reported | Pages 3-6 |  |  |
| Objectives | 3 | State specific objectives, including any prespecified hypotheses | Page 6 |  |  |
| **Methods** | | | | | |
| Study Design | 4 | Present key elements of study design early in the paper | Page 6 |  |  |
| Setting | 5 | Describe the setting, locations, and relevant dates, including  periods of recruitment, exposure, follow-up, and data collection | Page 6 |  |  |

| Participants | 6 | (a) *Cohort study* - Give the eligibility criteria, and the sources and methods of selection of participants. Describe methods of follow-up   *Case-control study* - Give the eligibility criteria, and the sources and methods of case ascertainment and control selection. Give the rationale for the choice of cases and controls *Cross-sectional study* - Give the eligibility criteria, and the sources and methods of selection of participants   (b) *Cohort study* - For matched studies, give matching criteria and number of exposed and unexposed   *Case-control study* - For matched studies, give matching criteria and the number of controls per case | Page 7 | RECORD 6.1: The methods of study population selection (such as codes or algorithms used to identify subjects) should be listed in detail. If this is not possible, an explanation should be provided.  RECORD 6.2: Any validation studies of the codes or algorithms used to select the population should be referenced. If validation was conducted for this study and not published elsewhere, detailed methods and results should be provided.  RECORD 6.3: If the study involved linkage of databases, consider use of a flow diagram or other graphical display to demonstrate the data linkage process, including the number of individuals with linked data at each stage. | Pages 6-7, supplementary table 2  Page 8  Page 6, supplementary table 1 |
| --- | --- | --- | --- | --- | --- |
| Variables | 7 | Clearly define all outcomes, exposures, predictors, potential confounders, and effect modifiers. Give diagnostic criteria, if applicable. | Page 7, Supplementary equation 1 | RECORD 7.1: A complete list of codes and algorithms used to classify exposures, outcomes, confounders, and effect modifiers should be provided. If these cannot be reported, an explanation should be provided. | Page 7, Supplementary equation 1 |
| Data sources/ measurement | 8 | For each variable of interest, give sources of data and details of methods of assessment (measurement).  Describe comparability of assessment methods if there is more than one group | Page 6, Supplementary table 1 |  |  |

| Bias | 9 | Describe any efforts to address potential sources of bias | Page 7 |  |  |
| --- | --- | --- | --- | --- | --- |
| Study size | 10 | Explain how the study size was arrived at | Page 6 |  |  |
| Quantitative variables | 11 | Explain how quantitative variables were handled in the analyses. If applicable, describe  which groupings were chosen, and why | Page 7, Supplementary equation 1 |  |  |
| Statistical methods | 12 | (a) Describe all statistical methods, including those used to control for confounding (b) Describe any methods used to examine subgroups and interactions (c) Explain how missing data were addressed (d) *Cohort study* - If applicable, explain how loss to follow-up was addressed   *Case-control study* - If applicable, explain how matching of cases and controls was addressed  *Cross-sectional study* - If applicable, describe analytical methods taking account of sampling strategy   (e) Describe any sensitivity analyses | Pages 7-8 |  |  |
| Data access and cleaning methods |  | .. | Pages 6-7 | RECORD 12.1: Authors should describe the extent to which the investigators had access to the database population used to create the study population. | Page 6 |

|  |  |  |  | RECORD 12.2: Authors should provide information on the data cleaning methods used in the study. | Pages 6-7 |
| --- | --- | --- | --- | --- | --- |
| Linkage |  | .. | Page 6 | RECORD 12.3: State whether the study included person-level, institutional-level, or other data linkage across two or more databases. The methods of linkage and methods of  linkage quality evaluation should be provided. | Page 6 |
| **Results** | | | | | |
| Participants | 13 | (a) Report the numbers of individuals at each stage of the study (*e.g.*, numbers potentially eligible, examined for eligibility, confirmed eligible, included in the study, completing follow-up, and analysed) (b) Give reasons for non- participation at each stage. (c) Consider use of a flow diagram | Page 6, table 1 (pages18-19) | RECORD 13.1: Describe in detail the selection of the persons included in the study (*i.e.,* study population selection) including filtering based on data quality, data availability and linkage. The selection of included persons can be described in the text and/or by means of the study flow diagram. | Page 6 |
| Descriptive data | 14 | (a) Give characteristics of study participants (*e.g.*, demographic, clinical, social) and information on exposures and potential confounders (b) Indicate the number of participants with missing data for each variable of interest (c) *Cohort study* - summarise follow-up time (*e.g.*, average and total amount) | Page 9, table 1  (pages 18-19),  supplmentary table 5 |  |  |
| Outcome data | 15 | *Cohort study* - Report numbers of outcome events or summary measures over time  *Case-control study* - Report numbers in each exposure | table 1  (pages 18-19),  supplmentary table 5 |  |  |

|  |  | category, or summary measures of exposure  *Cross-sectional study* - Report numbers of outcome events or summary measures |  |  |  |
| --- | --- | --- | --- | --- | --- |
| Main results | 16 | (a) Give unadjusted estimates and, if applicable, confounder- adjusted estimates and their precision (e.g., 95% confidence interval). Make clear which confounders were adjusted for and why they were included (b) Report category boundaries when continuous variables were categorized (c) If relevant, consider translating estimates of relative risk into absolute risk for a meaningful time period | Supplementary table 3, supplementary  equation 1 |  |  |
| Other analyses | 17 | Report other analyses done— e.g., analyses of subgroups and interactions, and sensitivity analyses | Page 10 |  |  |
| **Discussion** | | | | | |
| Key results | 18 | Summarise key results with  reference to study objectives | Pages 10-11 |  |  |
| Limitations | 19 | Discuss limitations of the study, taking into account sources of potential bias or imprecision.  Discuss both direction and magnitude of any potential bias | Page 11 | RECORD 19.1: Discuss the implications of using data that were not created or collected to answer the specific research question(s). Include discussion of misclassification bias, unmeasured confounding, missing data, and changing eligibility over time, as they pertain to the study being  reported. | Page 11 |
| Interpretation | 20 | Give a cautious overall interpretation of results considering objectives, | Page 11 |  |  |

|  |  | limitations, multiplicity of analyses, results from similar studies, and other relevant evidence |  |  |  |
| --- | --- | --- | --- | --- | --- |
| Generalisability | 21 | Discuss the generalisability (external validity) of the study results | Pages 11-12 |  |  |
| **Other Information** | | | | | |
| Funding | 22 | Give the source of funding and the role of the funders for the present study and, if applicable, for the original study on which the present article is based | Page 9 |  |  |
| Accessibility of protocol, raw data, and programming  code |  | .. | Page 13 | RECORD 22.1: Authors should provide information on how to access any supplemental information such as the study protocol, raw data, or  programming code. | Page 13, Supplementary materal 1 |

*Reference: Benchimol EI, Smeeth L, Guttmann A, Harron K, Moher D, Petersen I, Sørensen HT, von Elm E, Langan SM, the RECORD Working Committee. The REporting of studies Conducted using Observational Routinely-collected health Data (RECORD) Statement. *PLoS Medicine* 2015; in press.

*Checklist is protected under Creative Commons Attribution ([CC BY](http://creativecommons.org/licenses/by/4.0/)) license.

**Supplementary material 1**

**Statistical Analysis Plan for** Estimating Vaccine Effectiveness of the Maternal RSV Vaccine Against Severe Disease in Infants in Scotland, UK.

1. **Introduction**

Respiratory syncytial virus (RSV) is a major cause of respiratory illness worldwide, with four in five children having evidence of prior infection by age two [1, 2]. While most cases cause mild symptoms, infants under six months—especially those born prematurely—are at higher risk of severe RSV-related complications like bronchiolitis and pneumonia [3]. In Scotland, 1,104 infants under three months were hospitalised with RSV in 2023 (96 admissions per 1,000 infants) [4].

Recognising the potential for severe RSV disease in selected age groups, the Joint Committee on Vaccination and Immunisation (JCVI) recommended implementation of a vaccination programme to protect infants and older adults from severe disease cause by RSV [5]. Commencing August 2024, Scotland launched its maternal RSV vaccination programme, offering bivalent RSV prefusion F protein-based (RSVpreF) vaccine (Pfizer) in a range of vaccination settings, and older adult programme, offering vaccination to those aged 75 to 79 years. Pregnant women are eligible for maternal vaccination from 28 weeks gestation although some women will not take up the offer until later in their pregnancies. Previously published data from randomised control trials show high levels of vaccine efficacy against severe disease in the infant following maternal RSV vaccination [6,7].

This work looks to estimate vaccine effectiveness in a real-world setting using two methodologies: a nested case-control study as the primary methodology and a target trial emulation study as a supporting methodology. Both are valid methods to estimate vaccine effectiveness but have different strengths and potential biases. Consistent estimates of vaccine effectiveness across the two methods would increase confidence in the estimates but are not guaranteed and therefore the nested case-control design will be used as the primary methodology.

**Nested case-control design**

Nested case-control studies can be used to investigate the effectiveness of public health interventions, such as vaccination programmes. A nested case-control study draws cases and a specified number of controls, in which the outcome has not occurred by the time of the case outcome, from the full cohort population. Cases and controls are matched on variables such as age to ensure they come from the same underlying population. For this analysis the nested case-control design will match at the time of hospitalisation and look back to vaccination status.

**Target trial emulation design**

Target trial emulation is framework used in observational studies to design analysis as if a randomised trial were being run. It can be used to evaluate the effectiveness of interventions such as vaccination. To mimic randomisation propensity score matching is used to match vaccinated and unvaccinated mothers based on their propensity to be vaccinated. The propensity score for individuals is estimated based on observed characteristics. A target trial emulation design enables individuals to be followed up to explore RSV hospitalisation and other birth outcomes such as pre-term birth. This methodology is more complex than the nested case-control method due to the need to predict, with a high degree of accuracy, whether individuals (mothers) are likely to get vaccinated or not and changes to the delivery of the maternal RSV vaccine after an initial ‘catch-up’ programme.

2. **Study Objectives**

**Primary objective:** to estimate vaccine effectiveness of the maternal RSV vaccine against severe disease in infants.

3. **Study Population**

The study population comprises singleton pregnancies and resulting live births to women in Scotland with end of pregnancy date between 5 August 2024 and 31 December 2024. Between 2020 and 2024, there were an average of 46,650 live births per year in Scotland (NRS) [8]. Based on this figure, we expect a sample size of approximately 18,915 live births during the study period.

**Start date:** 5 August 2024 (earliest maternal RSV vaccination in the data).

**End date:** 31 December 2024 (main analysis), 31 March 2025 (secondary analysis). An end date of 31 December 2024 will cover the main RSV season (figure 1) while an end date of 31 March 2025 provides scope to look at vaccine waning.

4. **Descriptive Statistics**

Descriptive statistic will be employed to visually inspect differences in the proportion of and/or trends in vaccination uptake and RSV hospitalisations.

Plots:

- Births trend
- Maternal age distribution

Calculate the proportion (vaccination and RSV-related hospital admissions) and cumulative incidence (RSV-related hospital admissions) for the following variables:

- Baby sex
- Birthweight (re-coded: very low (<1500g), low (1500-2499g) and normal (>=2500g))
- Gestational age at birth (re-coded: very pre-term - <32 weeks, pre-term – 32-36 weeks, term – 37+ weeks)
- Maternal age at end of pregnancy (re-coded: <25, 25-29, 30-34, 35+ years)
- Maternal ethnicity
- Maternal smoking at antenatal booking
- Previous delivery (re-coded: 0, 1, 2+)
- SIMD (maternal SIMD at end of pregnancy)
- Total births this pregnancy

5. **Statistical software**

Analyses will be carried out using R/RStudio, version 4.4.2. Packages: Epi (version 2.59), ggsurvfit (version 1.1.0), survival (version 3.8.3), tidycmprsk (version 1.1.0).

6. **Variables/Data Sources**

Datasets will be accessed using through the Respiratory Data Platform. All datasets within the platform are linkable using the Community Health Index number (CHI number). The CHI number uniquely identifies a patient within the NHS in Scotland. The Respiratory Data Platform is a register of all patients in NHS Scotland which has been pseudonymised (respiratory ID, CHI numbers are used for data processing only and are not available for analysis), deduplicated and cleaned prior to being made available for analysis in a folder on the PHS stats server (access restricted to authorised PHS staff). ECOSS, SMR01, NRS deaths will be linked using baby’s respiratory ID and RSV vaccination data (using mother’s respiratory ID to create a cohort of live births with date of birth between 5 August 2024 and 31 December 2024. Variables to be used in both the nested case-control and target trial emulation designs are outlined in Table 1.

7. **Reporting results**

Results will be written in (a) manuscript(s) and submitted to a peer-review journal. Findings will also be presented at academic and clinical conferences, and with Scottish Government and other interested parties as appropriate.

Vaccine effectiveness estimates from the primary analysis (nested case-control design) will be published ahead of vaccination (in our respiratory report/a separate report/pre-print in July) for the next winter season.

**Nested case-control**

Using a nested case-control design cases will be matched to controls at the time of infant RSV hospitalisation, matching on gestational age at birth and ISO week of birth, and looking back to maternal RSV vaccination status.

1. **Study Objectives**

**Secondary objective:** to assess estimates of vaccine effectiveness by maternal age, maternal SIMD, maternal smoking, maternal ethnicity**,** number of previous deliveries, gestational age at birth, birthweight, baby’s sex (Table 1).

2. **Study Population**

**Inclusion criteria:**

- Singleton live births with end of pregnancy date (i.e., date of birth) between 5 August 2024 and 31 December 2024.

**Exclusion criteria:**

- Individuals part of multiple births. In the nested case-control design multiple births will be excluded to prevent overmatching (where a case and control is part of the same case-control set) and biases and confounding (due to shared genetics and environment).

**End points:**

- Death (baby)
- End of study (end date)
- RSV hospital admission (baby)
- Transferred out (GP) (baby)

3. **Outcome**

**Outcome of interest:** first RSV-related hospitalisation.

The first RSV-related hospitalisation will capture the earliest moment the vaccine might have failed to prevent severe disease, and subsequent hospitalisations could be biased by changes to healthcare seeking behaviour or other complications.

Any RSV-related hospitalisations will be defined as an emergency admission with a respiratory main diagnosis (J00-J99, U07.1 and U07.2) and an RSV positive PCR test 14 days before or within 2 days of hospital admission. Those with a positive RSV test but no hospital admission will not be considered as having the outcome of interest. This study outcome relates to severe illness - i.e. hospitalisation.

4. **Exposure**

**Exposure:** maternal RSV vaccination during pregnancy. Vaccination status will be categorised as:

- unvaccinated (no maternal RSV vaccine prior to the end of pregnancy)
- partially vaccinated (received maternal RSV vaccine <14 days before the end of pregnancy)
- vaccinated (received maternal RSV vaccine >= 14 days before the end of pregnancy)

5. **Covariates**

**Age:** maternal age at end of pregnancy (re-coded: <25, 25-29, 30-34, 35+ years)

**Birthweight:** birthweight (re-coded: very low (<1500g), low (1500-2499g) and normal (>=2500g))

**Ethnicity:** maternal ethnicity

**Previous delivery:** number of previous deliveries (re-coded: 0, 1, 2+)

**Sex:** baby’s sex

**SIMD:** maternal SIMD at the end of pregnancy

**Smoking:** maternal smoking at antenatal booking

6. **Method**

**Cases:** within a cohort of live births, delivered 5 August 2024 to 31 December 2024, any individual with an RSV-related hospital admission. Infants will be <=22 weeks olds at the end of the study period.

**Matching:** Cases will be matched to controls at the time of RSV-related hospitalisation looking back to vaccination status. Cases will be matched to 10 controls on ISO week of birth and gestational age at birth. A random selection of controls from the general population will be matched to each case using the ccwc() function in Epi, R. For pre-term babies, where weekly number of births are low, gestational age will be categorised as < 32 weeks gestation, 32-34 weeks, 35 & 36 weeks and in individual weeks from 37 weeks onward to increase the number of complete case-control sets.

We will use conditional logistic regression to estimate the odds ratio (OR) of maternal RSV vaccination using the following models:

- **Unadjusted:** $\boldsymbol{clogit(outcome\sim protection+strata(Set)}$
- **Adjusted:** $\boldsymbol{clogit(outcome\sim protection+age+birthweight+ethnicity+previous.delivery+sex+SIMD+smoking+strata(Set)}$

Where**:**

- **Protection:** vaccination status (unvaccinated, partially vaccinated (birth within 2 weeks of vaccination), vaccinated
- **Age:** maternal age at end of pregnancy (re-coded: <25, 25-29, 30-34, 35+ years)
- **Birthweight:** birthweight (re-coded: very low (<1500g), low (1500-2499g) and normal (>=2500g))
- **Ethnicity:** maternal ethnicity
- **Previous.delivery:** number of previous deliveries (re-coded: 0, 1, 2+)
- **Set:** case-control group
- **Sex:** baby’s sex
- **SIMD:** maternal SIMD at the end of pregnancy
- **Smoking:** maternal smoking at antenatal booking

Vaccine effectiveness (VE) will be estimated as:

VE = (1-OR)*100

**Subgroup analysis:** subgroup analysis by age at hospitalisation and gestational age at birth could be performed.

**Target trial emulation**

The target trial approach matches women vaccinated against RSV on a given date during pregnancy to other pregnant women, unvaccinated on same date, based on their propensity to be vaccinated (1:1 ratio). If the unvaccinated woman is vaccinated at a later date the set will be censored. Resulting live births will be followed up to observe the outcome (can also be used to look at baby outcomes e.g. pre-term birth).

1. **Study Objectives**

**Secondary objective:** to assess estimates of vaccine effectiveness by maternal age, maternal SIMD, maternal smoking, maternal ethnicity**,** number of previous deliveries, gestational age at birth, birthweight, baby’s sex (Table 1).

*Place / delivery (HB, urban-rurality, model, midwives numbers, variables from Tomi modelling work)*

*Seasonality (awareness campaigns, disease seasons)*

*Timing (gestation, calendar time, time since programme start)*

*Previous engagement with healthcare (history of covid variation etc)*

**Teritiary objective:** to assess baby outcomes (e.g. pre-term birth) following maternal RSV vaccination.

2. **Study Population**

**Inclusion criteria:**

- Pregnant women eligible for the maternal RSV vaccination between 5 August 2024 and 31 December 2024.

**Exclusion criteria:**

- Mothers with missing maternal age and unknown CHI.

**End points:**

- Death (baby)
- End of study (end date)
- RSV hospital admission (baby)
- Transferred out (GP) (baby)

3. **Outcome**

**Primary outcome of interest:** first RSV-related hospitalisation.

The first RSV-related hospitalisation will capture the earliest moment the vaccine might have failed to prevent severe disease, and subsequent hospitalisations could be biased by changes to healthcare seeking behaviour or other complications.

Any RSV-related hospitalisations will be defined as an emergency admission with a respiratory main diagnosis (J00-J99) and an RSV positive PCR test 14 days before or within 2 days of hospital admission. Those with a positive RSV test but no hospital admission will not be considered as having the outcome of interest. This study outcome relates to severe illness – i.e. hospitalisation.

**Secondary outcome of interest:** birth related outcomes (e.g. pre-term birth).

As individuals are followed up from the time of vaccination (time of matching) it is possible to explore baby related outcomes such as pre-term birth.

4. **Exposure**

**Exposure:** maternal RSV vaccination during pregnancy. Vaccination status will be categorised as:

- unvaccinated (no maternal RSV vaccine prior to the end of pregnancy)
- partially vaccinated (received maternal RSV vaccine <14 days before the end of pregnancy)
- vaccinated (received maternal RSV vaccine >= 14 days before the end of pregnancy)

5. **Covariates**

- **Estimated date of conception**
- **Maternal age at conception**
- **Maternal BMI**
- **Maternal country of birth group**
- **Maternal ethnicity**
- **Maternal SIMD at antenatal booking**
- **Maternal smoking**
- **Total foetuses this pregnancy**

6. **Method**

**Matching:** The variables outlined in Table 2 will be included in propensity matching of vaccinated:unvaccinated mothers. A 1:1 matching ratio will be used. Propensity matching matches individuals based on a single probability that summarises their likelihood to be vaccinated given a number of different covariates.

**Table 2: Factors to be included in propensity matching**

| **Category** | **Variable** | **Details/Notes** |
| --- | --- | --- |
| Maternal factors | Age |  |
|  | BMI |  |
|  | Comorbidities | Number of respiratory/cardiac comorbidities, asthma (/allergic conditions), diabetes, obesity |
|  | Deprivation | SIMD/other factors to describe this? |
|  | Ethnicity | Other factors to describe this? |
|  | Immunosuppression status |  |
|  | IVF |  |
|  | Number of previous children/birth order |  |
|  | Previous engagement with healthcare | History of COVID vaccinations, pertussis etc *COVID but not pertussis in the respiratory data platform* |
|  | Previous miscarriage | Is there reliable data? |
|  | Recent hospitalisation |  |
|  | Recent prescribing |  |
|  | Smoking |  |
| Place/delivery | Delivery model |  |
|  | Health Board |  |
|  | Midwife numbers |  |
|  | Urban-rurality |  |
|  | Variables from Tomi modelling work |  |
| Seasonality | Awareness campaigns |  |
|  | Disease seasons |  |
| Timing | Calendar time | Day granularity? |
|  | Gestation |  |
|  | Time since programme start |  |

**Appendix**

**Table 1: Grouping of variables by source**

|  | **Data Category** | **Data Item** | **Data Source** | **Design** |
| --- | --- | --- | --- | --- |
| Exposure | Vaccination | Delivery model | Vaccinations dataset/NCDS? | Target trial emulation |
|  |  | Health board | Vaccinations dataset/NCDS? | Target trial emulation |
|  |  | Midwife numbers |  | Target trial emulation |
|  |  | Urban-rurality |  | Target trial emulation |
|  |  | Vaccine | Vaccinations dataset/NCDS | Both |
|  |  | Vaccine date | Vaccinations dataset/NCDS | Both |
| Outcome | Laboratory testing | RSV PCR test result | ECOSS | Both |
|  |  | Specimen date | ECOSS | Both |
|  | Mortality | NRS death | NRS | Both |
|  |  | Infant death flag | SLiPBD | Both |
|  | Other | Transfer out | GP | Both |
|  | Secondary care | Admission date | SMR01 | Both |
|  |  | Emergency admission flag | SMR01 | Both |
|  |  | Level of care? *Is this variable in the respiratory data platform?* | SMR01 | Both |
|  |  | Lower Respiratory Tract Infection diagnosis | SMR01 | Nested case-control? |
|  |  | Main diagnosis (respiratory) | SMR01 | Nested case-control |
| Confounders/  covariates | Birth | Birthweight | SLiPBD | Nested case-control |
|  |  | End of pregnancy (date) | SLiPBD | Both |
|  |  | Gestational age | SLiPBD | Both |
|  |  | IVF |  | Target trial emulation |
|  |  | Outcome (live births) | SLiPBD | Both |
|  |  | Number births this pregnancy | SLiPBD | Both |
|  |  | Number previous deliveries | SLiPBD | Both |
|  | Seasonality/timing | Awareness campaigns |  | Both |
|  |  | Calendar time |  |  |
|  |  | Disease seasons |  |  |
|  |  | Time since programme start |  |  |
|  | Socio-demographics | Age (mother, at end of pregnancy) | SLiPBD | Both |
|  |  | Ethnicity (mother) | SLiPBD | Both |
|  |  | NHS health board of residence (at end of pregnancy)? | SLiPBD | Target trial emulation |
|  |  | Sex (baby) | SLiPBD | Nested case-control |
|  |  | SIMD (mother, at end of pregnancy) | SLiPBD | Both |
|  | Other | BMI (mother) |  | Target trial emulation |
|  |  | Comorbidities | GP? | Both |
|  |  | Immunosuppression status (baby? mother?) | GP? | Target trial emulation |
|  |  | Previous engagement with healthcare | Vaccinations dataset/NCDS? | Target trial emulation |
|  |  | Smoking (mother, at antenatal booking) | SLiPBD | Both |

Electronic Communication of Surveillance Scotland (ECOSS), GP records, Scottish Morbidity Records (SMR01), Scottish Linked Pregnancy and Baby Dataset (SLiPBD), National Records of Scotland (NRS), Vaccinations dataset: Data for vaccinations occurring in Scotland, and for Scottish residents vaccinated elsewhere is held in the National Clinical Data Store (NCDS)

Figure 1: RSV positivity in under 1s (from https://www.opendata.nhs.scot/dataset/viral-respiratory-diseases-including-influenza-and-covid-19-data-in-scotland/resource/378fb6c1-fc57-410a-94ff-a4c9722938c1)

RSV positivity rates in under 1s

| Week | Positivity rate per 100,000 |
| --- | --- |
| 20240701 | 0 |
| 20240708 | 6.5 |
| 20240715 | 8.7 |
| 20240722 | 6.5 |
| 20240729 | 2.2 |
| 20240805 | 13 |
| 20240812 | 6.5 |
| 20240819 | 8.7 |
| 20240826 | 8.7 |
| 20240902 | 21.6 |
| 20240909 | 28.1 |
| 20240916 | 19.5 |
| 20240923 | 23.8 |
| 20240930 | 41.1 |
| 20241007 | 52 |
| 20241014 | 75.8 |
| 20241021 | 134.2 |
| 20241028 | 114.7 |
| 20241104 | 199.1 |
| 20241111 | 272.7 |
| 20241118 | 339.8 |
| 20241125 | 261.9 |
| 20241202 | 339.8 |
| 20241209 | 251.1 |
| 20241216 | 270.6 |
| 20241223 | 212.1 |
| 20241230 | 214.3 |
| 20250106 | 97.4 |
| 20250113 | 93.1 |
| 20250120 | 64.9 |
| 20250127 | 67.1 |
| 20250203 | 34.6 |
| 20250210 | 49.8 |
| 20250217 | 28.1 |
| 20250224 | 28.1 |
| 20250303 | 19.5 |
| 20250310 | 26 |
| 20250317 | 13 |
| 20250324 | 21.6 |
| 20250331 | 10.8 |
| 20250407 | 6.5 |
| 20250414 | 10.8 |
| 20250421 | 4.3 |
| 20250428 | 6.5 |

Figure 2: Directed acyclic diagram

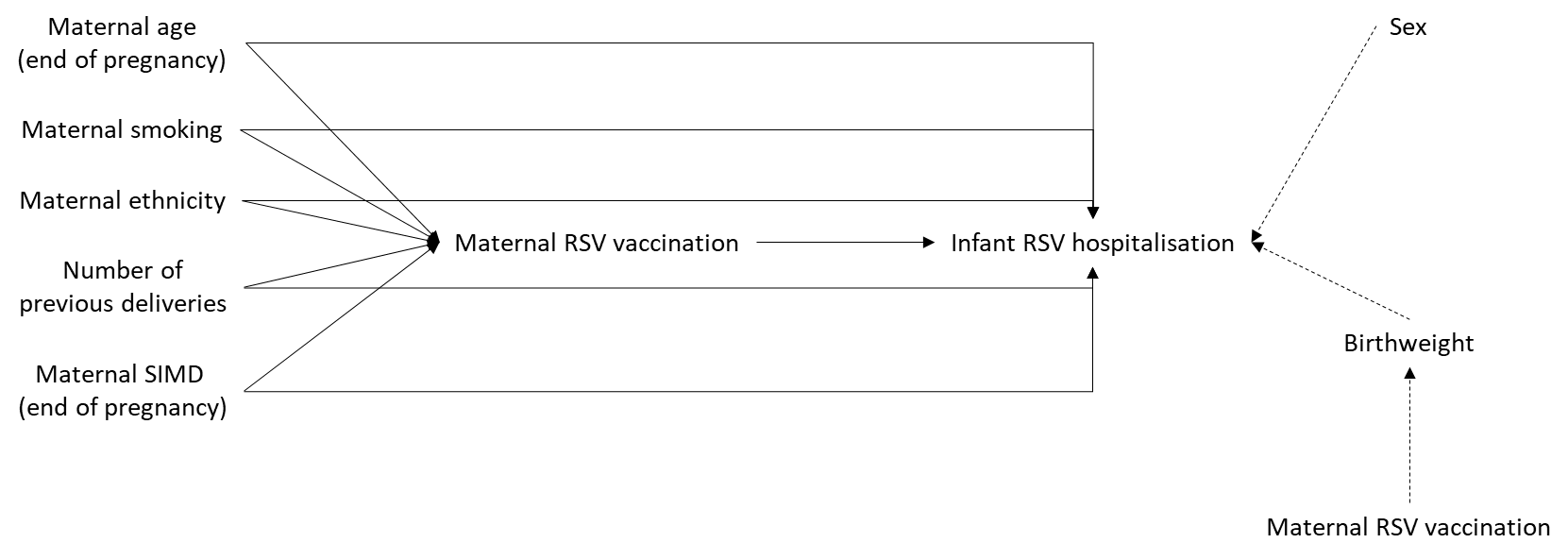

8.	[Births, deaths and other vital events - fourth quarter 2024 - National Records of Scotland (NRS)](https://www.nrscotland.gov.uk/publications/births-deaths-and-other-vital-events-fourth-quarter-2024/)
